# Molecular epidemiology of invasive Group A Streptococcal infections before and after the COVID-19 pandemic in Switzerland

**DOI:** 10.1101/2024.04.03.24305261

**Authors:** Angeliki M. Andrianaki, Jessica Franz, Federica Andreoni, Judith Bergada-Pijuan, Thomas C. Scheier, Tanja Duwe, Marc Pfister, Helena Seth-Smith, Tim Roloff, Natalia Kolesnik-Goldmann, Sara H. Burkhard, Alexia Cusini, Urs Karrer, Christian Rüegg, Adrian Schibli, Jacques Schrenzel, Stefano Musumeci, Roger D. Kouyos, Adrian Egli, Silvio D. Brugger, Annelies S. Zinkernagel

**Author notes:** Corresponding author: Prof. Dr. Dr. med. Annelies Zinkernagel Department of Infectious Diseases and Hospital Epidemiology University Hospital of Zurich, Raemistrasse 100, 8091 Zurich, Switzerland.

## Abstract

Group A *Streptococcus* (GAS, aka *Streptococcus pyogenes*) poses a significant public health concern, causing a diverse spectrum of infections with high mortality rates. Following the COVID-19 pandemic, a resurgence of invasive GAS (iGAS) infections has been documented, necessitating efficient outbreak detection methods. Whole genome sequencing (WGS) serves as the gold standard for GAS molecular typing, albeit constrained by time and costs. This study aimed to characterize the post-pandemic increased prevalence of iGAS on the molecular epidemiological level in order to assess whether new, more virulent variants have emerged, as well as to assess the performance of the rapid and cost-effective Fourier-transform infrared (FT-IR) spectroscopy as an alternative to WGS for detecting and characterizing GAS transmission routes. A total of 66 iGAS strains isolated from nine Swiss hospitals during the COVID-19 post-pandemic increased GAS prevalence were evaluated and compared to 15 strains collected before and 12 during the COVID-19 pandemic. FT-IR measurements and WGS were conducted for network analysis. Demographic, clinical, and epidemiological data were collected. Skin and soft tissue infection was the most common diagnosis, followed by primary bacteremia and pneumonia. Viral co-infections were found in 25% of cases and were significantly associated with more severe disease requiring intensive care unit admission. WGS analysis did not reveal emerging GAS genetic distinct variants after the COVID-19 pandemic, indicating the absence of a pandemic-induced shift. FT-IR spectroscopy exhibited limitations in differentiating genetically distant GAS strains, yielding poor overlap with WGS-derived clusters. The *emm1/*ST28 gebotype was predominant in our cohort and was associated with five of the seven deaths recorded, in accordance with the molecular epidemiological data before the pandemic. Additionally, no notable shift in antibiotic susceptibility patterns was observed. Our data suggest that mainly non-pathogen related factors contributed to the recent increased prevalence of iGAS.

## Introduction

Group A *Streptococcus* (GAS, aka *Streptococcus pyogenes*) is a pathobiont that causes a wide spectrum of infections, ranging from pharyngitis and superficial skin infections to severe invasive infections, such as necrotizing fasciitis, pneumonia or meningitis associated with high mortality and morbidity rates, with 500,000 estimated deaths annually worldwide [1–3]. After a historically low incidence of GAS infections during the COVID-19 pandemic, an alarmingly increasing incidence of invasive GAS (iGAS) infections has been reported, since October 2022, in many European countries and in the USA, mostly among children under 10 years of age, including many fatalities [4–7]. Also in Switzerland, since November 2022, a fourfold increase of the registered iGAS infections in children, including four deaths, has been reported compared to the pre-COVID-19 pandemic era [8]. So far, there is no evidence for the emergence of a more virulent clone [9, 10]. In the United Kingdom the pre-pandemic predominant M1uk clone was detected in the majority of the GAS strains isolated during the December 2022 outbreak [9]. The causes of the resurgence of iGAS remain somewhat elusive.

The detection and management of outbreaks, particularly in the case of GAS, rely on understanding transmission routes and genetic diversity. Molecular typing by whole genome sequencing (WGS) or conventional (Sanger) sequencing of the hypervariable N-terminal region of the *emm* gene, encoding the surface M protein [11], is crucial for epidemiological studies and outbreak investigations, although it is time-consuming, expensive and requires expertise [12, 13]. Alternatively, Fourier-transform infrared (FT-IR) spectroscopy may provide a cost-effective approach for outbreak analysis by generating a biochemical fingerprint of the bacterial composition using the full infrared spectrum (4000-400 cm^−1^), albeit with variable cluster defining cut-offs among different bacterial species and dependencies on growth conditions [13–19].

The aim of this study was to evaluate whether any GAS genomic variations had emerged. To this end, we compared iGAS strains isolated before, during, and after the COVID-19 pandemic (from January 2013 until May 2023) from patients hospitalized in nine hospitals in the Regions of Zurich, Graubünden and Geneva using the current gold standard WGS. Furthermore, we aimed to assess whether FT-IR spectroscopy is a reliable alternative for a time-efficient detection of GAS outbreaks compared to WGS.

## Materials and Methods

### Study period, population, clinical and epidemiological information

The study encompassed 93 strains sourced from the same number of patients with GAS infection. Among these, 15 strains were retrospectively identified between January 2013 and December 2019, 18 strains were retrieved from the COVID-19 pandemic period between February 2020 and September 2022. The remaining 60 strains were prospectively identified during the latest iGAS resurgence (October 2022 to May 2023) from patients hospitalized in nine Swiss hospitals: University Hospital Zurich, University Hospital of Geneva, City Hospital of Zurich, Cantonal Hospital of Graubünden, Cantonal Hospital of Winterthur, Regional Hospitals of Wetzikon, Uster, Limmatal, and Männedorf. These hospitals collectively serve an area inhabited by approximately 2 million individuals. Only three of the participating hospitals admitted pediatric patients. Two patients did not have an invasive infection (GAS_036 and GAS_037), but were included in the study because they were related to a patient who died from iGAS infection (GAS_035). Basic demographical, clinical, and epidemiological data were collected for 74 patients from electronic health records, including sex, age, diagnosis, co-infections, sampling date and site, as well as the geographical location of the sampling. Outcomes included admission to the intensive care unit (ICU), intubation and death related to iGAS. No clinical data were available from the Regional Hospitals of Limmatal and Männedorf.

In order to determine the prevalence of GAS over the years, the number of all GAS strains detected by culture or molecular techniques in patients admitted to the University Hospital of Zurich between January 2012 and December 2023 was collected. The GAS strains originating from blood, joint punctures, cerebrospinal fluid, bronchoalveolar lavage, pleura fluid and soft tissue were defined as invasive. The definition of the Centre for Disease Control regarding invasive infection was adopted [20].

### Ethics approval

This study was approved by the Ethic Committee of Nordwest- and Central Switzerland with the agreement of all Swiss Ethical Committees (BASEC-ID: 2019-01291 as part of the Swiss Pathogen Surveillance Platform, www.spsp.ch). Patients declining to sign a general consent or any other declining statement against using data for research purposes were excluded.

### Bacterial strains

93 GAS strains were collected from the same number of patients hospitalized in the nine hospitals mentioned above and stored at -80°C. Species identification of all samples was carried out using Matrix Assisted Laser-Desorption Ionization – Time of Flight (MALDI-TOF) (MBT Compass 4.1, Bruker Daltonics, Bremen, Germany) using the database BDAL DB.13.

### Sample preparation for FT-IR spectroscopy and WGS

Three to five macroscopically identical colonies were subcultured on 5 % sheep blood agar plates (Biomérieux) for 24±2 hours at 37°C in a static incubator in the presence of 5% CO_2_. For FT-IR spectroscopy, a 1μl-loop was overloaded with bacteria cells that were then resuspended in 50 μl ethanol solution (70% v/v) in a 1.5 ml tube containing metal rods. The suspension was then vortexed extensively and 50 μl of deionized water were added, followed by another round of vortexing. Fifteen microliters of the homogenized bacterial suspension were placed on a 96-spot silicon plate (Bruker Daltonics, Bremen, Germany) in four technical replicates per bacterial isolate. The plates were dried at 37°C for approximately 20 minutes followed by infrared measurements. For WGS, a lawn was streaked from frozen stocks and samples were processed as previously described [21, 22].

### Infrared measurements

An IR-Biotyper (Bruker Daltonics) was used for all measurements according to the manufacturer’s instructions and as previously described [19]. In brief, 12 µl of test standard 1 and 2 (ITRS 1/ ITRS 2) were placed on the 96-spot silica plate and used as controls. The samples were measured using the OPUS software v8.2.28 (Bruker Daltonics), detecting carbohydrates spectra within the wavenumber range of 1200 and 900 cm^-1^. Data analysis was performed using the same software. Isolates with three or more valid replicates in the same run were included in the analysis.

### Whole genome sequencing of isolates and comparative genomics

Whole genome sequencing of the clinical isolates was performed at the Institute for Medical Microbiology, University of Zurich, Switzerland as previously described [21]. *Emm*-typing was extrapolated from the WGS data using the *emm*-typing tool [23] and MLST typing was inferred from the WGS data using MLST v.2.7.6 [24], which makes use of the PubMLST website (https://pubmlst.org/) developed by Keith Jolley [25] and sited at the University of Oxford. The development of that website was funded by the Wellcome Trust.

De novo assemblies were built with SPAdes v.3.14.1 using the –careful option [26]. Annotation of the resulting assemblies was performed with Prokka v1.14.6 [27]. The created GFF files were used as input to Roary v3.13.0 to construct the pangenome, for which we used the command -e -mafft that generates a multi-fasta alignment of the core genes [28]. The average nucleotide identity (ANI) was inferred with pyani v0.2.12 choosing the method ANIm, which makes use of MUMmer / NUCmer to align the input sequences [29]. Phylogenetic trees were inferred from the aligned core genome using IQTREE [30] under the GTR+G+I model. Trees and metadata were processed and plotted using the R-packages ape [31] and ggtree [32].

The overlap between clusters derived from FT-IR and whole bacterial genome data (specifically Average Nucleotide Identity, ANI) was quantified as outlined in Scheier *et al* [19] . In brief FT-IR clusters were inferred by cutting the hierarchical (UPGMA) clustering tree derived from the Euclidean distance of the FT-IR spectra at a given height, which was varied over a broad range. ANI-based clusters were defined as the components of the network obtained by connecting those pairs of nodes whose ANI was above a given threshold. We used the v-measure to quantify the overlap between these two networks.

The genomic data with matched phenotypic information can be downloaded here (National Center for Biotechnology Information (nih.gov)).

### Antibiotic susceptibility testing

Antibiotic susceptibility testing to penicillin, clindamycin and erythromycin was conducted on pandemic and post-pandemic GAS strains as part of the routine microbiological assessment. The Kirby-Bauer method was perfomed in line with the recommendations of the European Committee on Antimicrobial Susceptibility Testing (EUCAST) in the ISO accredited Institute of Molecular Microbiology (IMM) in Zurich. Briefly, a 0.5 McFarland standard inoculum preparation was used. Mueller-Hinton agar was supplemented with 5% defibrinated horse blood and 20 mg/L β-NAD (MH-F agar). After evenly inoculating the agar with the bacterial suspension, antimicrobial disks were applied to the agar surface. To detect inducible clindamycin resistance, erythromycin and clindamycin disks were placed 12-16 mm apart (edge to edge). The plates were incubated at 35 ± 1°C in with 4-6% CO_2_ for 18 ± 2 hours, after which zones of inhibition were measured to determine susceptibility based on EUCAST breakpoint tables (version 14.0).

### Data Analysis

For the patients’ clinical characteristics we performed a descriptive statistical analysis. Continuous variables were presented as mean with range, except for age which was presented as median with range. Categorical variables were presented as frequency tables. For the association of the patient-related factors with the severity of the disease we chose as dependent variables the ICU admission and the all-cause in-hospital mortality and performed a univariable analysis. Due to the rare event problem for the case of obesity (4/74 incidences), we collapsed this variable with two other related medical conditions, diabetes mellitus and cardiovascular disease (CVD), based on their clinical relevance and exhibited overlap. Variables that demonstrated significant (or marginally significant) associations (p<0.07) in the univariable analysis were taken to multivariable binary logistic regression analysis. The significance level was set at p<0.05. The model fit was evaluated using the Hosmer-Lemeshow test. All analyses were performed with IBM SPSS Statistics V26 (IBM Corp., Armonk, NY, USA).

### Funding information

This study was supported by the University Of Zurich CRPP Personalized Medicine Of Persisting Bacterial Infections Aiming to Optimize Treatment and Outcome to S.D.B, and A.S.Z.; and by grant (INOV00121) from University Hospital Zurich to T.C.S.

## Results

### Report of laboratory-confirmed GAS cases at the University Hospital Zurich between January 2012 and December 2023

The total number of iGAS strains detected with cultures or molecular techniques from January 2012 to December 2023 reported at the University Hospital Zurich is shown in **Figure 1**. GAS infections typically have a seasonal pattern, showing a peak during late spring in the northern hemisphere [33]. During the COVID-19 pandemic (from March 2020 until the suspension of the pandemic prevention methods in Switzerland, in March 2022), the detection of GAS cases declined compared to the previous years (**Suppl. Figure 1**) and the number of iGAS was also historically low (**Figure 1**). In the first quarter of 2023, there was a notable surge in invasive and non-invasive GAS cases, marking a threefold increase compared to the same month average in pre-pandemic years (**Figure 1, Suppl. Figure 1).** The number of isolated GAS strains remained unusually high throughout the first post-pandemic year.

**Figure 1.**
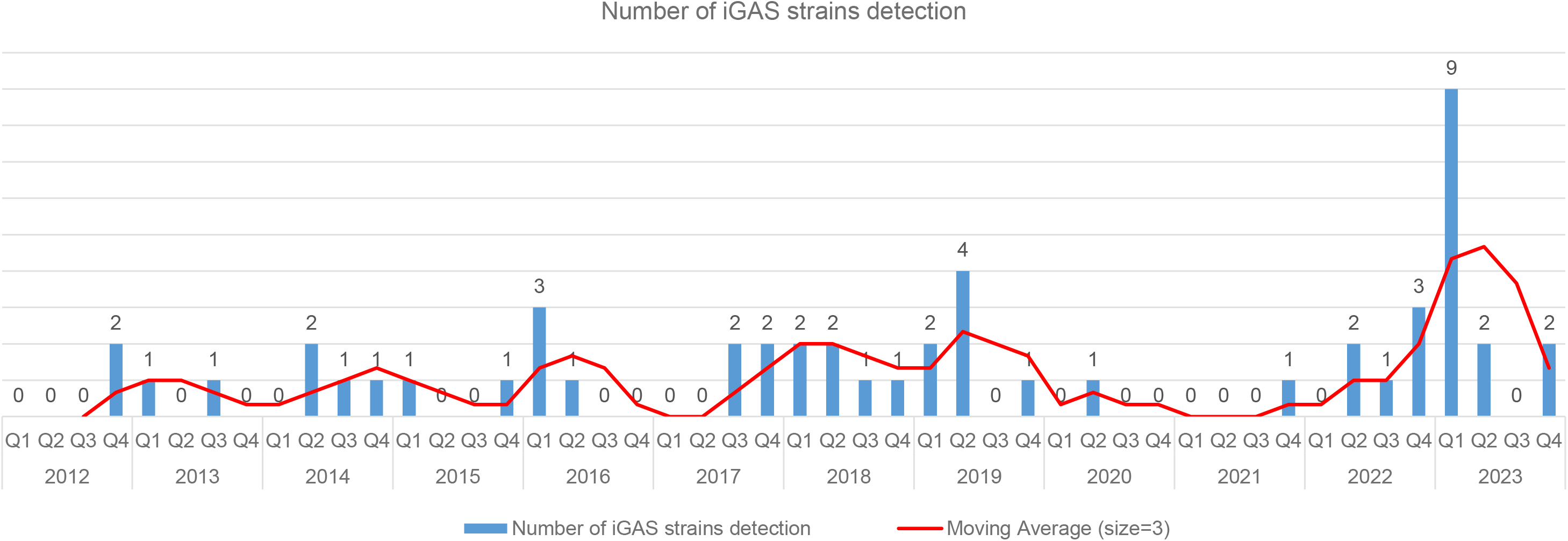
Number of detected invasive GAS strains at the University Hospital Zurich between January 2012 and December 2023. Invasive GAS (iGAS) strains detected either by culturing or via molecular methods are included in this graph. The blue color indicates the number of new cases monthly and the red color the moving average every three months. Q1: months from January to March, Q2: months from April to June, Q3: months from July to September, Q4: months from October to December.

### Demographic and clinical characteristics of the patients with iGAS infections

93 patients, from whom the 93 GAS strains were isolated, were included in this study. The clinical data of 74 patients hospitalized in seven Swiss hospitals were included. The clinical data of the remaining 19 patients (14 from the retrospectively collected strains and five from the prospectively collected ones) could not be retrieved. 44 patients (59.5%) were male with a median age of 47.5 years (range 1 to 94 years), nine (6.5%) were under 18 years of age. Cardiovascular disease (35.1%), diabetes mellitus (17.6%), and solid tumors (9.5%) were the most common comorbidities. The most common diagnosis of infection was skin and soft tissue infection (SSTI) with bacteremia (18.9%), whereas necrotizing fasciitis was diagnosed in five (6.8%) patients. The primary sampling site, representing the majority of cases (64.9%), was blood, followed by tissue biopsies (13.5%). Notably, 24.3% of cases featured viral co-infections; six patients (8.2%) were diagnosed with influenza and three (4.1%) with SARS-CoV-2, three (4.1%) with a co-infection with SARS-CoV-2 and influenza and three (4.1%) with respiratory syncytial virus (RSV). Two patients were diagnosed with varicella zoster virus (VZV) reactivation. Fifteen patients (20.3%) suffered from toxic shock syndrome (TSS). A large number required ICU admission (43.2%) and intubation (29.7%). Surgical debridement was performed in 39 patients (52.7%). Combination antimicrobial therapy was employed in 38 (51.4%) of cases; in 27 of them (71%) clindamycin was co-administered with another antibiotic. Immunoglobulin was administered to all fifteen patients with TSS (20.3%). The average hospitalization duration was 15.6 days, (range 0 - 100 days). We observed an all-cause mortality rate of 9.5%, with 6 out of 7 deaths specifically attributed to iGAS. The demographic and clinical characteristics of the patients are summarized in **Table 1**.

**Table 1.** Demographic and clinical characteristics of the patients with invasive GAS infections.

|  |  |
| --- | --- |
| **Characteristics (N=74)** |  |
| **Sex, n (%)** |  |
| Male | 44 (59.5) |
| Female | 30 (40.5) |
| **Median age (range), years** | 47.5 (1-94) |
| **Patients < 18 years old, n (%)** | 9 (6.5) |
| **Hospital, n (%)** |  |
| University Hospital of Zurich | 20 (27) |
| University Hospital of Geneva | 17 (23) |
| City Hospital Zurich | 16 (21.6) |
| Wetzikon Hospital | 9 (12.2) |
| Cantonal Hospital Chur | 7 (9.5) |
| Uster Hospital | 4 (5.4) |
| Cantonal Hospital Winterthur | 1 (1.4) |
| **Coexisting conditions, n (%)** |  |
| Cardiovascular disease | 26 (35.1) |
| Diabetes mellitus | 13 (17.6) |
| Renal disease | 8 (10.8) |
| Solid tumortumour | 7 (9.5) |
| Alcohol abuse | 6 (8.1) |
| Intravenous drug use | 5 (6.8) |
| Immunosuppression | 4 (5.4) |
| Adipositas | 4 (5.4) |
| Chronic obstructive lung disease | 3 (4.1) |
| Autoimmune disease | 2 (2.7) |
| Viral hepatitis | 2 (2.7) |
| HIV | 1 (1.4) |
| HematologicalHaematological malignancy | 0 (0) |
| **No underlying disease, n (%)** | 30 (40.5) |
| **≥ 1 underlying disease, n (%)** | 44 (59.5) |
| **Homeless** | 2 (2.7) |
| **Sampling site, n (%)** |  |
| Blood | 48 (64.9) |
| Tissue biopsy | 10 (13.5) |
| Swab | 5 (6.8) |
| Joint fluid | 3 (4.1) |
| Pleural fluid | 3 (4.1) |
| Abscess | 3 (4.1) |
| Bone biopsy | 1 (1.4) |
| Ascites | 1 (1.4) |
| **Diagnosis, n (%)** |  |
| SSTI with bacteraemia | 14 (18.9) |
| Pneumonia | 12 (16.2) |
| Isolated bacteraemia | 10 (13.5) |
| SSTI without bacteraemia | 5 (6.8) |
| Necrotizing fasciitis | 5 (6.8) |
| Septic arthritis | 5 (6.8) |
| Non pregnancy related endometritis | 3 (4.1) |
| Osteomyelitis | 3 (4.1) |
| Tonsilitis with abscess | 3 (4.1) |
| Septic bursitis | 3 (4.1) |
| Otitis media/mastoiditis without bacteraemia | 2 (2.7) |
| Otitis media/mastoiditis with bacteraemia | 2 (2.7) |
| Tonsilitis without abscess | 2 (2.7) |
| Pharyngitis with bacteraemia | 1 (1.4) |
| Meningitis | 1 (1.4) |
| Pregnancy related endometritis | 1 (1.4) |
| Mediastinitis | 1 (1.4) |
| Endocarditis | 1 (1.4) |
| **Toxic shock syndrome, n (%)** | 15 (20.3) |
| **Septic shock, n (%)** | 32 (43.5) |
| **Viral co-infection, n (%)** | 18 (24.3) |
| Influenza (A or B) | 6 (8.2) |
| SARS-CoV-2 | 3 (4.1) |
| SARS-CoV-2 and Influenza (A or B) | 3 (4.1) |
| RSV | 3 (4.1) |
| VZV | 2 (2.7) |
| Rhinovirus | 1 (1.4) |
| **ICU admission, n (%)** | 32 (43.2) |
| **Intubation, n (%)** | 22 (29.7) |
| **Surgery, n (%)** | 39 (52.7) |
| **Amputation, n (%)** | 4 (5.4) |
| **Antimicrobial therapy, n (%)** |  |
| Co-Amoxicillin-Clavulanate | 39 (52.7) |
| Clindamycin | 36 (48.6) |
| CeftriaxonCeftriaxone | 26 (35.1) |
| Penicillin | 21(28.4) |
| Piperacillin-Tazobactam | 15 (20.3) |
| Vancomycin | 9 (12.2) |
| Meropenem | 4 (5.4) |
| Clarithromycin | 3 (4.1) |
| Cefepime | 3 (4.1) |
| Gentamycin | 2 (2.7) |
| **Combination therapy, n (%)** | 38 (51.4) |
| **Immunoglobulin, n (%)** | 15 (20.3) |
| **Duration of hospitalization, mean (range), days** | 15.6 (0-100) |
| **Duration of ICU hospitalization, mean (range), days** | 3.2 (0-21) |
| **All-cause mortality, n (%)** | 7 (9.5) |
| **GAS-related death, n (%)** | 6 (8.1) |

HIV: Human Immunodeficiency Virus, SSTI: Skin and soft tissue infections, ICU: Intensive Care Unit, GAS: Group A Streptococci, SARS-CoV-2: Severe acute respiratory syndrome coronavirus type 2, RSV: Respiratory Syncytial Virus, VZV: Varicella zoster virus

The univariable logistic regression analysis revealed significant correlations between ICU admission and viral coinfections (OR 3.600, 95% CI 1.172-11.057). A marginally significant correlation between ICU admission, female sex (OR 0.396, 95% CI 0.152-1.027), and the collapsed variable of obesity, cardiovascular disease and diabetes mellitus (OR 2.500, 95% CI 0.954-6.552) was detected **(Table 2).** All four patients with obesity were admitted to the ICU (p-Fisher 0.03). No significant correlations were found between the investigated patient-related variables and the all-cause in-hospital mortality **(Suppl. Table 1).** A multivariable bimodal logistic regression model for ICU admission confirmed statistically significant coefficients for female sex (OR 0.256, 95% CI 0.081 - 0.804), viral coinfection (OR 4.094, 95%CI 1.199-13.986) and the collapsed variable (obesity, cardiovascular disease or diabetes mellitus) (OR 4.595, 95% CI 1.424-14.835) **(Table 2).** The Nagelkerke R square was 0.256 indicating an adequate level of explanatory power and the significance level of the Hosmer-Lemeshow test was 0.393 indicating a good model fitting.

**Table 2.** Results of the univariable and multivariable logistic regression analysis of the association of patient-related factors with ICU admission (n=32) upon iGAS infection.

|  | Odds Ratio | 95% Confidence Intervall | p value |
| --- | --- | --- | --- |
| ***Univariable analysis*** |  |  |  |
| Female sex | 0.396 | 0.152, 1.027 | 0.057 |
| Cardiovascular disease | 1.944 | 0.739, 5.116 | 0.178 |
| COPD | 0.645 | 0.056, 7.445 | 0.725 |
| Diabetes mellitus | 1.680 | 0.504, 5.600 | 0.398 |
| Solid tumour | 0.194 | 0.022, 1.696 | 0.138 |
| Immunosuppression | 0.419 | 0.042, 4.232 | 0.461 |
| Alcohol abuse | 1.345 | 0.253, 7.150 | 0.728 |
| Renal disease | 0.766 | 0.169, 3.471 | 0.729 |
| Viral coinfection | 3.600 | 1.172, 11.057 | 0.062 |
| Collapsed metabolic Variable | 2.500 | 0.954, 6.552 |  |
| ***Multivariable analysis*** |  |  |  |
| Sex | 0.256 | 0.081, 0.804 | 0.020 |
| Collapsed Metabolic Variable | 4.595 | 1.424, 14.835 | 0.011 |
| Viral coinfection | 4.094 | 1.199, 13.986 | 0.025 |
| Constant | 0.651 |  | 0.336 |

COPD: chronic obstructive pulmonary disease, HIV: human immunodeficiency virus, IBD: inflammatory bowel disease, Collapsed Metabolic Variable: cardiovascular disease, diabetes mellitus and obesity

### Clinical isolates characteristics

Clinical isolates were genetically characterized to determine their ST and *emm*-type (**Table 3**). The *emm* typing yielded successful results in 92 out of 93 strains, detecting 18 different *emm* types. Among these, *emm1-*type (*emm1.0, emm 1.2, emm1.3, emm1.6, emm1.7*) emerged as the predominant type in 63 cases (68%), followed by *emm12 (emm12.0, emm12.2, emm12.4)* in 7 cases (7.6%). The *emm1*-type/ST-28 was prevalent, and associated with higher mortality, since it was detected in 5/7 (71%) patients who died.

**Table 3.** Molecular and microbiological characteristics of the 93 GAS clinical isolates.

| Nr. | Strain ID | Lab ID | Isolation year | ST | emm type | Inhibition zone (mm) |  |  |
| --- | --- | --- | --- | --- | --- | --- | --- | --- |
|  |  |  |  |  |  | PEN | ERM | CLI |
| 1 | GAS_001 | 23021066 | 2022 | 28 | 1.0 | 27 | 26 | 23 |
| 2 | GAS_002 | 23021067 | 2023 | 39 | 4.0 | 22 | 22 | 19 |
| 3 | GAS_003 | 23021068 | 2023 | 28 | 1.3 | 29 | 27 | 23 |
| 4 | GAS_004 | 23021069 | 2023 | 28 | 1.0 | 26 | 24 | 18 |
| 5 | GAS_005 | 23021070 | 2023 | 28 | 1.3 | 30 | 28 | 23 |
| 6 | GAS_006 | 23021071 | 2023 | 28 | 1.3 | 34 | 31 | 26 |
| 7 | GAS_007 | CI1224 | 2017 | 28 | NA | NA | NA | NA |
| 8 | GAS_008 | CI1271 | 2017 | 28 | 1.0 | NA | NA | NA |
| 9 | GAS_009 | CI1296 | 2017 | 28 | 1.0 | NA | NA | NA |
| 10 | GAS_010 | CI1316 | 2017 | 28 | 1.0 | NA | NA | NA |
| 11 | GAS_011 | CI1453 | 2018 | 28 | 1.0 | NA | NA | NA |
| 12 | GAS_012 | CI1800 | 2019 | 1138 | 1.0 | 32 | 29 | 23 |
| 13 | GAS_013 | CI2261 | 2019 | 101 | 1.0 | NA | NA | NA |
| 14 | GAS_014 | CI348 | 2013 | 28 | 1.0 | NA | NA | NA |
| 15 | GAS_015 | CI350 | 2013 | 28 | 1.0 | NA | NA | NA |
| 16 | GAS_016 | CI407 | 2013 | 52 | 28 | NA | NA | NA |
| 17 | GAS_017 | CI4711 | 2020 | 63 | 77.0 | 30 | 12 | 25 |
| 18 | GAS_018 | CI4839 | 2020 | 28 | 1.0 | 33 | 31 | 25 |
| 19 | GAS_019 | CI5359 | 2020 | 59 | 8.0 | 37 | 32 | 27 |
| 20 | GAS_020 | CI543 | 2013 | 28 | 1.0 | NA | NA | NA |
| 21 | GAS_021 | CI6416 | 2020 | 267 | 1.0 | 24 | 22 | 20 |
| 22 | GAS_022 | CI655 | 2014 | NA | 28 | NA | NA | NA |
| 23 | GAS_023 | CI729 | 2015 | 167 | 118 | NA | NA | NA |
| 24 | GAS_024 | CI7790 | 2021 | 403 | 11.1 | 31 | 6 | 9 |
| 25 | GAS_025 | CI780 | 2015 | 167 | 118 | NA | NA | NA |
| 26 | GAS_026 | CI781 | 2015 | 101 | 89 | NA | NA | NA |
| 27 | GAS_027 | CI8226 | 2023 | 242 | 12.4 | 30 | 28 | 22 |
| 28 | GAS_028 | CI8227 | 2023 | 28 | 1.3 | 32 | 29 | 23 |
| 29 | GAS_029 | CI8236 | 2023 | 28 | 1.0 | 25 | 24 | 19 |
| 30 | GAS_030 | CI8237 | 2023 | 28 | 1.0 | 32 | 28 | 22 |
| 31 | GAS_031 | CI8239 | 2023 | 242 | 12.4 | 33 | 30 | 24 |
| 32 | GAS_032 | CI8240 | 2023 | 242 | 1.3 | 30 | 28 | 22 |
| 33 | GAS_033 | CI8241 | 2023 | 28 | 1.3 | 31 | 28 | 23 |
| 34 | GAS_034 | CI8242 | 2023 | 28 | 1.0 | 32 | 29 | 23 |
| 35 | GAS_035 | CI8243 | 2023 | 28 | 1.0 | 33 | 30 | 24 |
| 36 | GAS_036 | CI8244 | 2023 | 28 | 1.0 | 33 | 29 | 24 |
| 37 | GAS_037 | CI8245 | 2023 | 28 | 1.0 | 31 | 28 | 23 |
| 38 | GAS_038 | CI8248 | 2023 | 28 | 1.0 | 31 | 29 | 23 |
| 39 | GAS_039 | CI8249 | 2023 | 28 | 1.0 | 27 | 26 | 22 |
| 40 | GAS_040 | CI8252 | 2022 | 28 | 1.0 | 30 | 29 | 23 |
| 41 | GAS_041 | CI8253 | 2022 | 28 | 1.0 | 31 | 29 | 23 |
| 42 | GAS_042 | CI8254 | 2022 | 28 | 1.0 | 31 | 29 | 23 |
| 43 | GAS_043 | CI8255 | 2023 | 28 | 1.0 | 32 | 29 | 23 |
| 44 | GAS_044 | CI8256 | 2023 | 28 | 1.0 | 33 | 28 | 22 |
| 45 | GAS_045 | CI8257 | 2023 | 28 | 1.0 | 31 | 29 | 23 |
| 46 | GAS_046 | CI8258 | 2023 | 36 | 12 | 31 | 28 | 23 |
| 47 | GAS_047 | CI8259 | 2023 | 28 | 1.0 | 32 | 28 | 23 |
| 48 | GAS_048 | CI8260 | 2023 | 36 | 12 | 32 | 28 | 23 |
| 49 | GAS_049 | CI8261 | 2023 | 62 | 1.0 | 34 | 30 | 24 |
| 50 | GAS_050 | CI8262 | 2023 | 28 | 1.0 | 29 | 28 | 22 |
| 51 | GAS_051 | CI8263 | 2023 | 28 | 1.0 | 32 | 29 | 23 |
| 52 | GAS_052 | CI8265 | 2023 | 36 | 12 | 32 | 28 | 23 |
| 53 | GAS_053 | CI8266 | 2023 | 46 | 22 | 35 | 14 | 31 |
| 54 | GAS_054 | CI8267 | 2022 | 39 | 1.0 | 26 | 26 | 21 |
| 55 | GAS_055 | CI8268 | 2022 | 28 | 1.0 | 24 | 26 | 22 |
| 56 | GAS_056 | CI8269 | 2023 | 28 | 1.0 | 24 | 22 | 18 |
| 57 | GAS_057 | CI8270 | 2023 | 28 | 1.0 | 26 | 22 | 19 |
| 58 | GAS_058 | CI8273 | 2023 | 148 | 1.3 | 29 | 26 | 21 |
| 59 | GAS_059 | CI8274 | 2023 | 28 | 1.3 | 33 | 28 | 23 |
| 60 | GAS_060 | CI8277 | 2023 | 28 | 1.0 | 28 | 24 | 19 |
| 61 | GAS_061 | CI8278 | 2023 | 28 | 1.0 | 22 | 23 | 18 |
| 62 | GAS_062 | CI8279 | 2022 | 28 | 1.0 | 31 | 30 | 19 |
| 63 | GAS_063 | CI8280 | 2023 | 314 | 1.0 | 25 | 12 | 17 |
| 64 | GAS_064 | CI8453 | 2022 | 28 | 1.0 | 32 | 29 | 23 |
| 65 | GAS_065 | CI8466 | 2022 | 101 | 1.0 | 27 | 25 | 19 |
| 66 | GAS_066 | CI8467 | 2022 | 28 | 1.0 | 33 | 28 | 22 |
| 67 | GAS_067 | CI9366 | 2023 | 28 | 1.4 | 29 | 25 | 22 |
| 68 | GAS_068 | CI9367 | 2023 | 101 | 89.4 | 27 | 28 | 22 |
| 69 | GAS_069 | CI9368 | 2023 | 62 | 87.9 | 26 | 23 | 21 |
| 70 | GAS_070 | CI9370 | 2023 | 242 | 12.4 | 24 | 22 | 22 |
| 71 | GAS_071 | CI9375 | 2022 | 28 | 1.7 | 27 | 24 | 19 |
| 72 | GAS_072 | CI9376 | 2022 | 28 | 1.4 | 27 | 24 | 21 |
| 73 | GAS_073 | CI9377 | 2022 | 28 | 1.7 | 34 | 32 | 30 |
| 74 | GAS_074 | CI9378 | 2022 | 178 | 44.0 | 24 | 24 | 20 |
| 75 | GAS_075 | CI9379 | 2022 | 190 | 49.3 | 28 | 28 | 22 |
| 76 | GAS_076 | CI9380 | 2022 | 36 | 12.2 | 19 | 22 | 18 |
| 77 | GAS_077 | CI9381 | 2022 | NA | 73.0 | 26 | 26 | 23 |
| 78 | GAS_078 | CI9382 | 2022 | NA | 80.0 | 30 | 30 | 27 |
| 79 | GAS_079 | CI9384 | 2022 | 137 | 104.0 | 33 | 30 | 25 |
| 80 | GAS_080 | CI9385 | 2022 | 190 | 49.3 | 27 | 26 | 21 |
| 81 | GAS_081 | CI9387 | 2022 | 28 | 1.4 | 31 | 31 | 24 |
| 82 | GAS_082 | CI9389 | 2022 | 242 | 252.0 | 26 | 24 | 22 |
| 83 | GAS_083 | CI9390 | 2022 | 150 | 212.0 | 26 | 26 | 22 |
| 84 | GAS_084 | FF09716211 | 2021 | 772 | 89.0 | 35 | 26 | 25 |
| 85 | GAS_085 | FS42596242 | 2022 | 11 | 53.0 | 30 | 26 | 25 |
| 86 | GAS_086 | FS49301899 | 2022 | 242 | 1.0 | 28 | 23 | 21 |
| 87 | GAS_087 | FS49818997 | 2022 | 28 | 1.0 | 32 | 33 | 24 |
| 88 | GAS_088 | FS55545334 | 2022 | 969 | 1.0 and<br>86.2 | 22 | 28 | 20 |
| 89 | GAS_089 | FS69217489 | 2023 | 28 | 1.0 | 27 | 24 | 22 |
| **90** | GAS_090 | FS72033123 | 2023 | 28 | 1.0 | 28 | 29 | 22 |
| **91** | GAS_091 | FS72033369 | 2023 | 28 | 1.0 | 30 | 24 | 22 |
| **92** | GAS_092 | FS72034424 | 2023 | 52 | 1.0 | 27 | 24 | 21 |
| **93** | GAS_093 | FS72034435 | 2023 | 28 | 1.0 | 29 | 24 | 20 |

NA: not available

The Kirby-Bauer method was performed on 79 strains in order to determine phenotypical antimicrobial resistance (**Table 3**). All tested strains were susceptible to penicillin. Only one strain was found resistant to clindamycin and erythromycin. No inducible clindamycin resistance was detected.

### Molecular typing of iGAS

A comprehensive genomic analysis utilizing WGS was performed on a cohort comprising 91 iGAS und 2 non-invasive GAS strains, whose sampling periods spanned through before, during, and after the COVID-19 pandemic **(Figure 2A and 2B).** Notably, we found a consistent preservation of sequence-types (STs) across all sampling times, indicating genetic stability within the GAS population. Intriguingly, most strains associated with the current GAS upsurge were found to originate from existing genetic lineages circulating before the pandemic, suggesting a continuum in transmission dynamics rather than a pandemic-induced shift. The phylogenies did not exhibit large clusters of recent and post-pandemic strains clearly distinct from the older reference variants refM1uk, ref5005, and ref5448. Ten strains (GAS_086, GAS_032, GAS_031, GAS_082, GAS_070, GAS_027, GAS_052, GAS_046,

**Figure 2.**
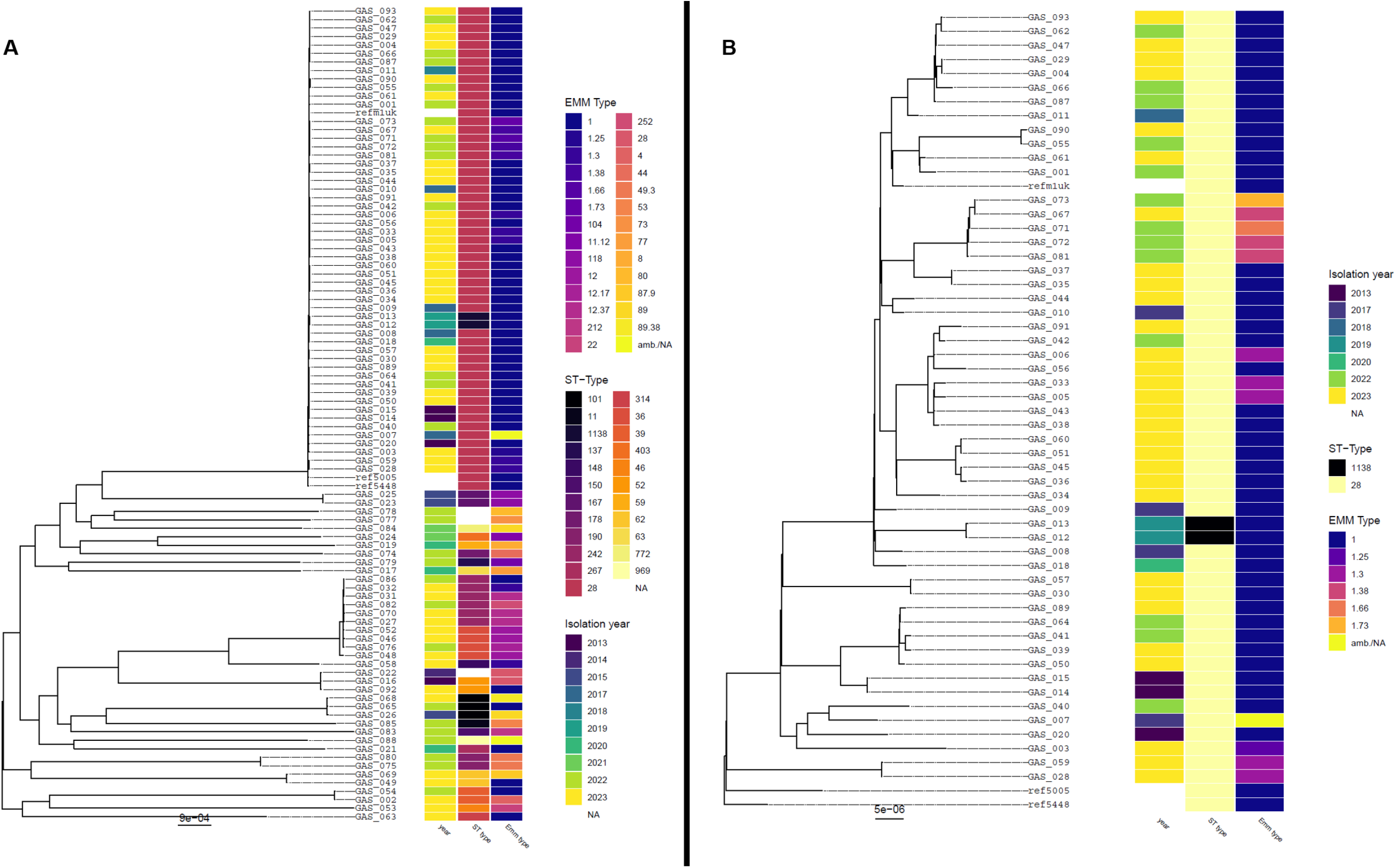
Phylogenetic tree of the 93 GAS clinical isolates and three reference strains (refM1uk, ref5005 and ref5448) Midpoint rooted maximum likelihood phylogenetic trees based on the alignment of the clinical isolates and reference strains core genes. The scale bar corresponds to the number of nucleotide substitutions per position. **A**. Phylogenic tree of all GAS clinical isolates and the three reference strains (refM1uk, ref5005 and ref5448). **B**. Magnified representation of the large *emm1*/ST28 cluster (including refM1uk, ref5005 and ref5448) from panel A.

GAS_076, GAS_048) exhibited a genetic resemblance by core genome phylogeny, possessing different ST types compared to the predominant ST-28 type. These strains were not confined to a specific geographic location but were dispersed throughout the country. A high level of genetic relatedness was found between GAS_035 and GAS_037 strains, originating from a patient that died from an iGAS infection and an individual related to the patient, respectively. FT-IR spectroscopy showed a poor concordance compared to the core genome analysis performed with the WGS, failing to reliably differentiate genetically distant GAS strains **(Fig. Suppl. 2)**.

## Discussion

In summary, this multicenter study shows that WGS core genome analysis of GAS strains isolated in Switzerland before, during, and after the COVID-19 pandemic did not reveal genetically distinct, emerging variants. The *emm1* ST28 was predominat in our cohort. FT-IR spectroscopy showed poor overlap with WGS and did not effectively differentiate genetically distant GAS strains, confirming WGS as the current gold standard for the characterization of GAS clinical isolates. Moreover, viral coinfections, mainly influenza and SARS-CoV-2, were significantly associated with a severe iGAS infection requiring an ICU admission.

While we found a large *emm1*/ST28 cluster, including the reference strains (refMIuk, ref5005 and ref5448), the absence of outbreak characteristics, such as clustered cases in a specific geographical region and a high genetic relatedness within potential transmission chains, suggests that the strains of the current resurgence were part of the diversity already circulating before the COVID-19 pandemic. The *emm1*/ST28 genotype has been commonly reported causing iGAS infections [34], consistently indicating its dominance over the years [9, 10, 35, 36]. Accordingly, we observed that in our cohort *emm1*/ST28 was the predominant variant, present in most patients (5/7) who died from an iGAS infection. Interestingly, we did not find any clusters originating from the three reference strains refM1uk, ref5005 and ref5448, known to cause invasive infections [9, 37, 38]. Furthermore, the phenotypic susceptibility testing revealed a high susceptibility to penicillin, erythromycin and clindamycin, confirming the known susceptibility patterns over the years without a shift towards more resistant strains, as reported worldwide [4, 6, 9].

The COVID-19 pandemic highlighted the need for rapid epidemic diagnosis in controlling outbreaks. To our knowledge, this study is the first to compare the time- and cost-effective FT-IR spectroscopy with WGS for GAS typing and outbreak investigation. FT-IR spectroscopy showed poor overlap with WGS and did not effectively differentiate genetically distant GAS strains. GAS, known for its adaptive nature and highly recombining genome, undergoes phenotypic changes during invasive infections, potentially impacting FT-IR spectroscopy’s efficacy, a technique reliant on biochemical fingerprints [39, 40]. Our findings underscore FT-IR spectroscopy’s limitations in characterizing GAS strains during outbreaks, supporting WGS as the gold standard typing method for GAS. Furthermore, WGS is a valuable tool for GAS characterization, since it can provide important information on *emm* type, virulence factors and antibiotic resistance genes.

Our epidemiological records from multiple centers across Switzerland confirm the dramatic increase in iGAS infections, also documented across numerous countries globally [4]. Even though these infections result in about 500,000 deaths annually worldwide, imposing a substantial economic burden [11, 41], data on the incidence of iGAS clinical manifestations remain scarce. At the University Hospital of Zurich, a threefold increase of the number of invasive and non-invasive GAS strains was reported since December 2022 compared to the pre-pandemic years, which aligns with the reports of other countries [6, 9]. Within our study, the most prevalent diagnosis was SSTI, in line with findings from a study conducted in New Zealand [42], followed by primary bacteremia and pneumonia. In accordance with other studies [43], viral coinfections, mostly influenza and SARS-CoV-2, were relatively frequent in our study cohort and significantly associated with a severe disease leading to ICU admission, supporting the current hypothesis linking the recent iGAS infections outbreak to a high prevalence of circulating respiratory viruses [10, 35]. Furthermore, we found that female sex and the metabolic risk factors (cardiovascular disease, obesity and diabetes mellitus) were also significantly associated with a severe disease requiring an ICU support. The all-cause mortality in our cohort was 9.5%, comparable with the data published in England [35], highlighting the severity of iGAS infections.

The cause of the global post-pandemic increased GAS infections prevalence remains elusive. Our findings point out that the recent upsurge does not arise from pathogen-related factors, prompting further exploration of alternative causes, as proposed in prior studies [6, 9, 10, 44].

The current hypothesis suggests that the lack of exposure to GAS and common seasonal respiratory viruses during the COVID-19 pandemic, due to isolation measures, resulted in decreased immunity to GAS and seasonal respiratory viruses [35, 37, 43, 45]. After lifting the COVID-19 pandemic protection measures, an increase and a shift of the seasonal pattern in seasonal respiratory viral and iGAS infections was observed after two years of their historically low incidence.

This study has some limitations that affect its broader applicability. The sample origin, limited to specific Swiss regions, may not represent a broader GAS strain diversity. The retrospectively collected subset of data and the unavailability of clinical information for some patients does not allow determining risk factors associated with the current increased iGAS prevalence. Only three of the participating hospitals admitted pediatric patients, leading to an underrepresentation of minors. To draw more decisive conclusions regarding the risk factors of iGAS infections, future studies should include larger, diverse samples cohorts from various regions, encompassing both invasive and non-invasive strains and performing genotypic as well as phenotypic virulence studies.

To the best of our knowledge, this is the biggest data set of invasive GAS strains across multiple centers in Switzerland, giving a broad view of the post-pandemic increased prevalence of iGAS infections. The long time-span of the examined GAS strains allowed us to explore potential genetic changes before and after the COVID-19 pandemic. The absence of genetic diversity and shift to more resistant strains throughout the years suggests that non-pathogen related factors may lead to resurgence of iGAS, supporting the theory that the lack of immunity to GAS and the common respiratory viruses due to the social distancing during the COVID-19 pandemic could be associated with severe iGAS infections with many fatalities.

## Conclusion

In summary, our study aimed to investigate the genetic characteristics of GAS strains responsible for the current resurgence of iGAS infection following the COVID-19 pandemic. WGS-based molecular typing did not uncover genetic changes regarding *emm*-type and sequence-type (ST) pre- during- and post-pandemic Moreover, antibiotic susceptibility testing revealed unchanged susceptibility patterns. The FT-IR spectroscopy limitations in GAS typing underscore the ongoing importance of WGS in molecular analysis. Our results suggest that non-pathogen related factors, such as the lack of immunity to GAS and other common respiratory viruses due to social distancing and wearing masks during the COVID-19 pandemic, may be responsible for the post-pandemic increased prevalence of the iGAS infections. Further research is needed to improve surveillance methods of iGAS strains.

**Supplementary Figure 1 Number of detected GAS strains (invasive and non-invasive) at the University Hospital Zurich between January 2012 and December 2023**

Invasive and non-invasive GAS strains detected either by culturing or via molecular methods are included in this graph. The blue color indicates the number of new cases monthly and the red color the moving average every three months. Q1: months from January to March, Q2: months from April to June, Q3: months from July to September, Q4: months from October to December.

**Supplementary Figure 2 Assessment of average nucleotide identity (ANI) networks with FT-IR spectroscopy**

Overlap (quantified as the v-measure) between FT-IR and ANI-derived clusters for different cutoffs used for FT-IR (X-axis) and ANI (color) clustering.

**Supplementary Table 1.** Results of the univariable and multivariable logistic regression analysis of the association of patient-related factors with all-cause in-hospital mortality (n=7) upon iGAS infection.

## Data Availability

All data produced in the present work are contained in the manuscript or available online at https://www.ncbi.nlm.nih.gov/

