## Supplementary figures and images for "Molecular epidemiology of invasive Group A Streptococcal infections before and after the COVID-19 pandemic in Switzerland"

### Supplementary figure 1

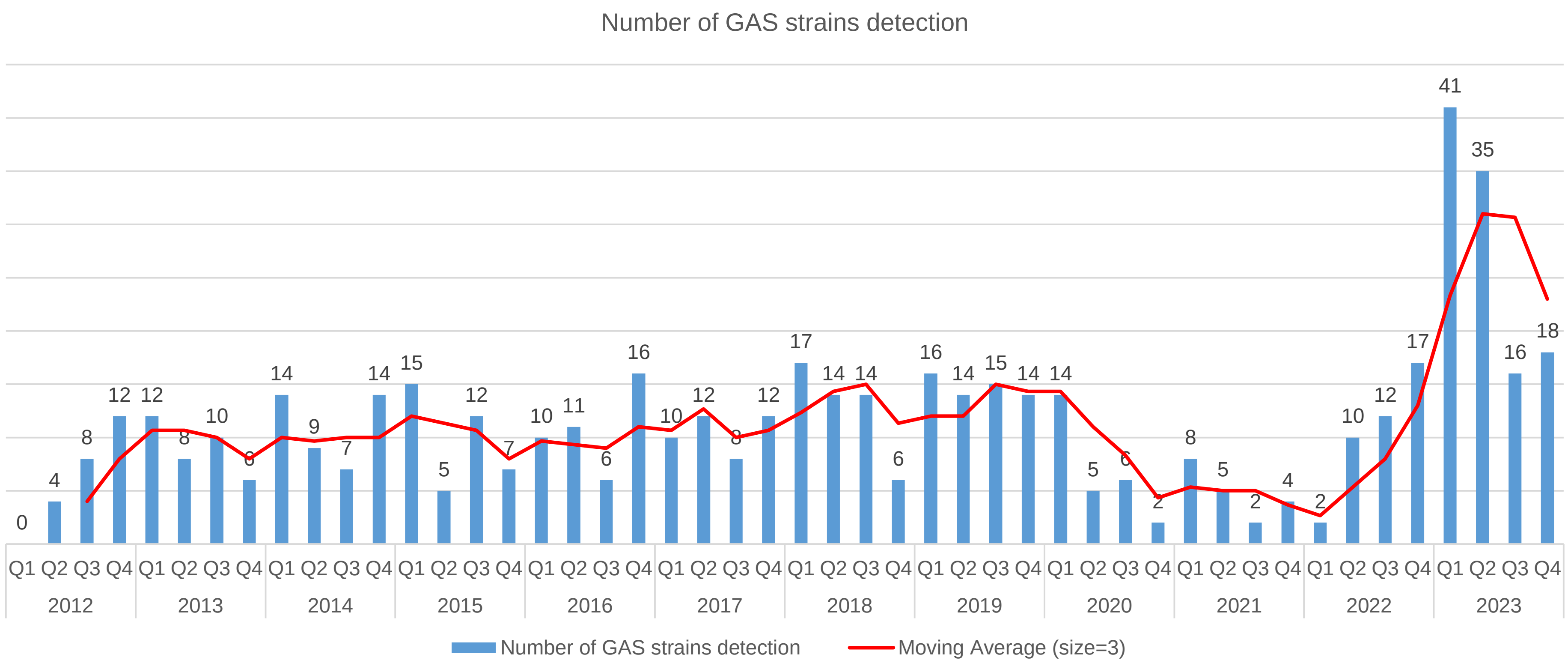

### Supplementary figure 2

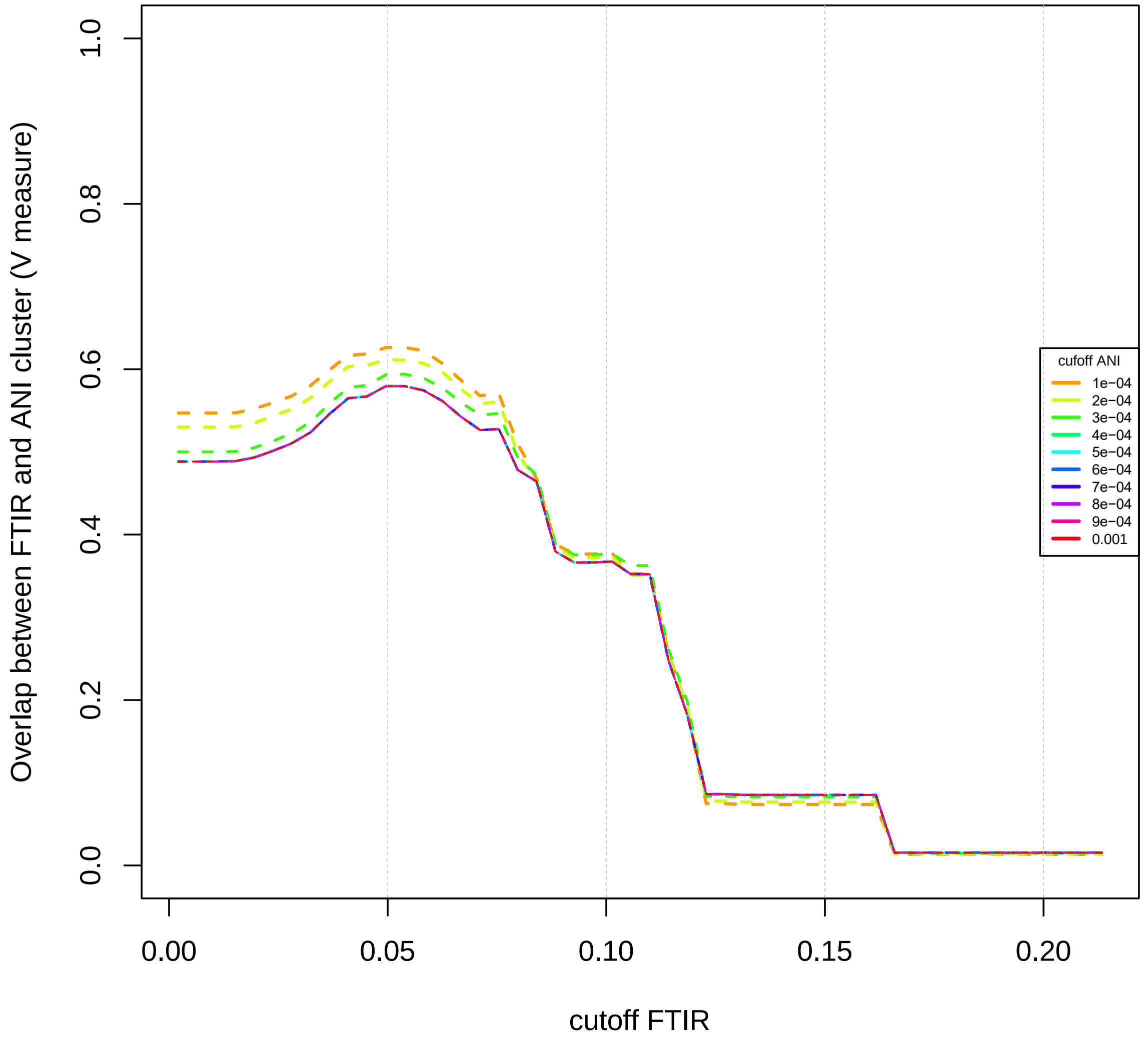
