## Supplementary table 1 for "Molecular epidemiology of invasive Group A Streptococcal infections before and after the COVID-19 pandemic in Switzerland"

|  | p value | Odds Ratio | 95% Confidence Intervall |
|---|---|---|---|
| Sex | 0.503 | 1.795 | 0.325, 9.926 |
| Cardiovascular disease | 0.214 | 2.727 | 0.561, 13.259 |
| COPD | 0.193 | 5.417 | 0.426, 68.816 |
| Diabetes melitus | 0.083 | 4.275 | 0.829, 22.057 |
| Solid tumor | 0.094 | 4.960 | 0.760, 32.375 |
| Immunosuppression | 0.303 | 3.556 | 0.318, 39.703 |
| Alcohol abuse | 0.999 | 0.000 | 0.000 |
| Renal disease | 0.135 | 4.067 | 0.645, 25.652 |
| Obesity | 0.999 | 0.000 | 0.000 |
| Viral Co-infection | 0.243 | 2.600 | 0.523, 12.921 |
| Collapsed Metabolic Variable | 0.074 | 4.783 | 0.860, 26.592 |
| ***Multivariable analysis*** | | | |
| Solid tumor | 0.313 | 2.796 | 0.380, 20.572 |
| Collapsed Metabolic Variable | 0.153 | 3.753 | 0.613, 22.982 |
| Constant | 0.000 | 0.044 | |

COPD: chronic obstructive pulmonary disease, HIV: human immunodeficiency virus, IBD: inflammatory bowel disease, Collapsed Metabolic Variable: cardiovascular disease, diabetes mellitus or obesity
